# Association of Microalbuminuria with High HbA_1_C levels in Melanesian Adults with Diabetes of at-least 1 Year Duration

**DOI:** 10.1101/2021.05.20.21257555

**Authors:** Ian Billy, Izzard Aglua

## Abstract

**Background:** Evidence suggest a potential relationship between high or variable HbA_1_C levels and presence or rate of change of microalbuminuria. Disruption of the vascular endothelial glycocalyx has been linked to chronic hyperglycemia and microalbuminuria, suggesting a possible shared pathophysiological mechanism.

**Aim:** To 1) explore potential association between microalbuminuria and high HbA_1_C levels in Melanesian adults with diabetes mellitus, and 2) asses predictive value of a high HbA_1_C reading as an indicator of the presence or progression of microalbuminuria.

**Method:** A cross-sectional study on 190 patients with either type 1 or 2 diabetes of at least 1-year duration done at a provincial hospital in Papua New Guinea in 2017.

**Result:** A significant [P=0.0221], though weak [R^2^=0.028 <0.70], correlation between UACR and HbA_1_C [95% statistical confidence] was observed in univariate regression, which was marginally significant [P=0.056] after controlling for weight, systolic hypertension, duration of diabetes, gender and age in multivariate regression.

**Conclusion:** A significant [p=0.022], though weak, correlation between UACR and HbA_1_C levels was observed, which may support usefulness of HbA_1_C as a predictor for microalbuminuria and diabetic kidney disease. In settings without microalbuminuria testing, high HbA1c levels can be used as a proxy to indicate presence or progression of microalbuminuria, thus prompting timely interventions to prevent further progression of diabetic nephropathy.

## Introduction

An estimated 40% of patients with type 1 or type 2 diabetes develop diabetic nephropathy within 5 to 15 years of diagnosis^1^. Microalbuminuria, defined as urine albumin of 30-300mg/day, is a useful clinical test for assessment of renal functional status or progressive deterioration in diabetes of many years^2,3^. Microalbuminuria testing has been widely accepted as an independent tool for early detection and monitoring of the presence or progression of insidious diabetic nephropathy to overt diabetic kidney disease^3,4^. Hemoglobin A_1_C (HbA_1_C) is a form of glycosylated hemoglobin resulting from non-enzymatic glycosylation of hemoglobin following longstanding hyperglycemia^5^. It serves as a reliable measure of glycemic control over a 90-120 days period, coinciding with erythrocyte life span^5,6^. Increasing evidence suggest a potential relationship between high or variable A_1_C levels and presence or rate of change of microalbuminuria^7,8^. Disruption of the vascular endothelial glycocalyx has been identified as a contributing mechanism to the development of microalbuminuria in diabetic kidney disease^9-12^. Chronic hyperglycemia has also been identified to independently contribute to systemic endothelial dysfunction, including endothelial glycocalyx dysregulation^12-14^. This suggests a potential biochemical association between chronic hyperglycemia and microalbuminuria.

This study was therefore done to 1) explore for a potential association between microalbuminuria (urine albumin-creatinine ratio>30) and high HbA_1_C readings (HbA_1_C≥6%) in Melanesian adults with diabetes of at least one year duration, and 2) explore the value of a high A_1_C reading (HbA_1_C≥6%), initial or progressive, at at-least 1 year after diagnosis, as an indicator for the presence or progression of microalbuminuria.

Most primary health care facilities in developing countries do not have point of care tests or laboratories for measuring microalbuminuria to inform treatment decisions^14^. Dipstick, a bedside test commonly available in most resource-constrained settings, can only detect overt albuminuria (≥ 300mg/day), and not microalbuminuria (30-300mg/day)^2^. Therefore, outcomes from this study are anticipated to alert treating doctors to the possible presence of microalbuminuria when encountering high or variable A_1_C levels, so that early and appropriate interventions can be instituted to minimize onset of subclinical microalbuminuria and its progression to macro albuminuria (proteinuria) or established diabetic nephropathy, as well as preventing related micro-or macro vascular diabetic complications^15,16^.

### Aim

- Explore for a potential association between microalbuminuria (urine albumin creatinine ratio >30 mg/day) and high HbA_1_C readings (HbA_1_C≥6%) in Melanesian adults with diabetes of at least one-year duration, and
- Explore the value of a high A_1_C reading (HbA_1_C≥6%), initial or progressive, at at-least 1 year after diagnosis as a reliable indicator of presence or progression of microalbuminuria.

## Method

A cross-sectional observational study on 190 Melanesian adults (age >13) with either type 1 or 2 diabetes of at least one-year duration was done at the Angau Memorial Hospital in Papua New Guinea in 2017, whereby single HbA_1_C and Urine Albumin-Creatinine Ratio (UACR) levels were measured in patients during one of their visits to the diabetic clinic. Venous blood was sampled from patients at the diabetic clinic by the treating doctor and sent in standard serum bottles for analysis at the hospital laboratory. HbA_1_C was measured using the SD-HbA1c analyzer while microalbuminuria was measured using the urine albumin-creatinine ratio from standard biochemical analyzers. Measurements were recorded on Microsoft Excel spreadsheet and analyzed using *Stata* statistical software. Linear regression models (univariate, multivariate) were performed on the data set for associations between UACR and HbA_1_C, gender, weight, age, systolic blood pressure and duration of diabetes. Patients with previous or co-existing other-cause-kidney-disease were not included in the study.

### Data and Resource Availability Statements

The datasets generated and/or analyzed during the current study are available from the corresponding author upon reasonable request. No applicable resources were generated or analyzed during the current study.

## Results

**Figure 1.**
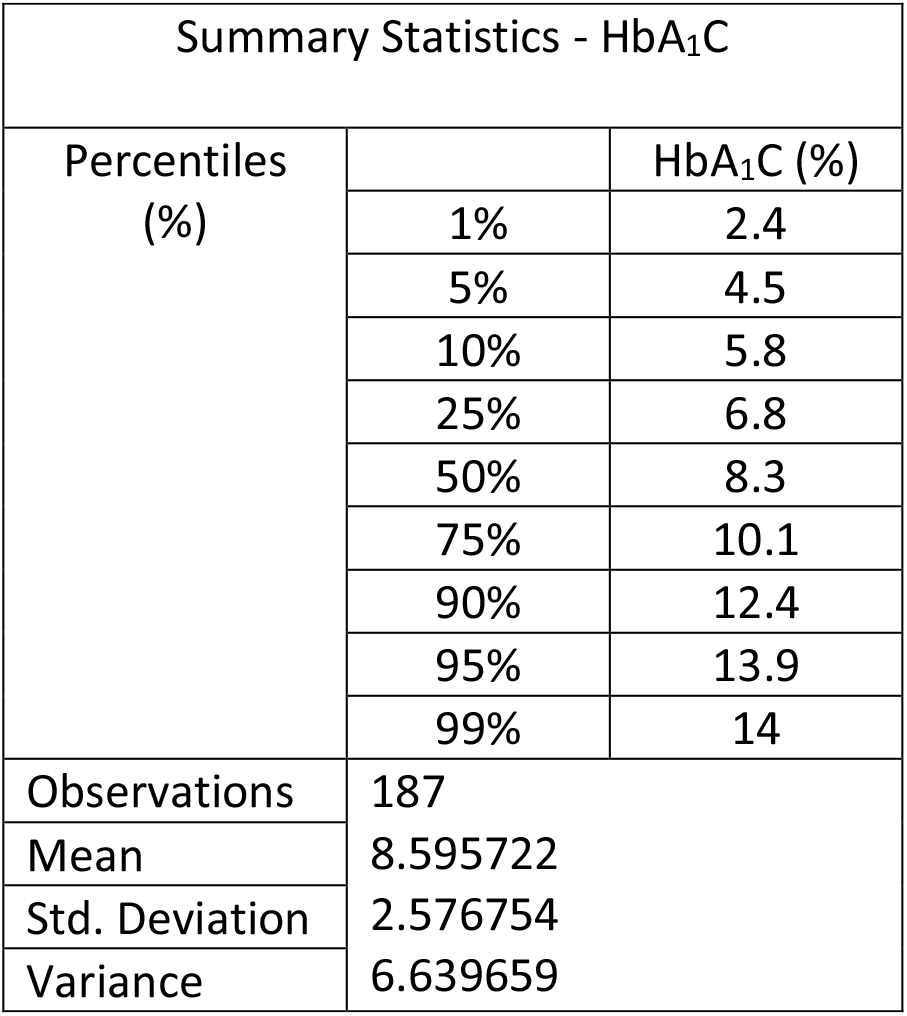
Summary statistics - HbA_1_C.

**Figure 2.**
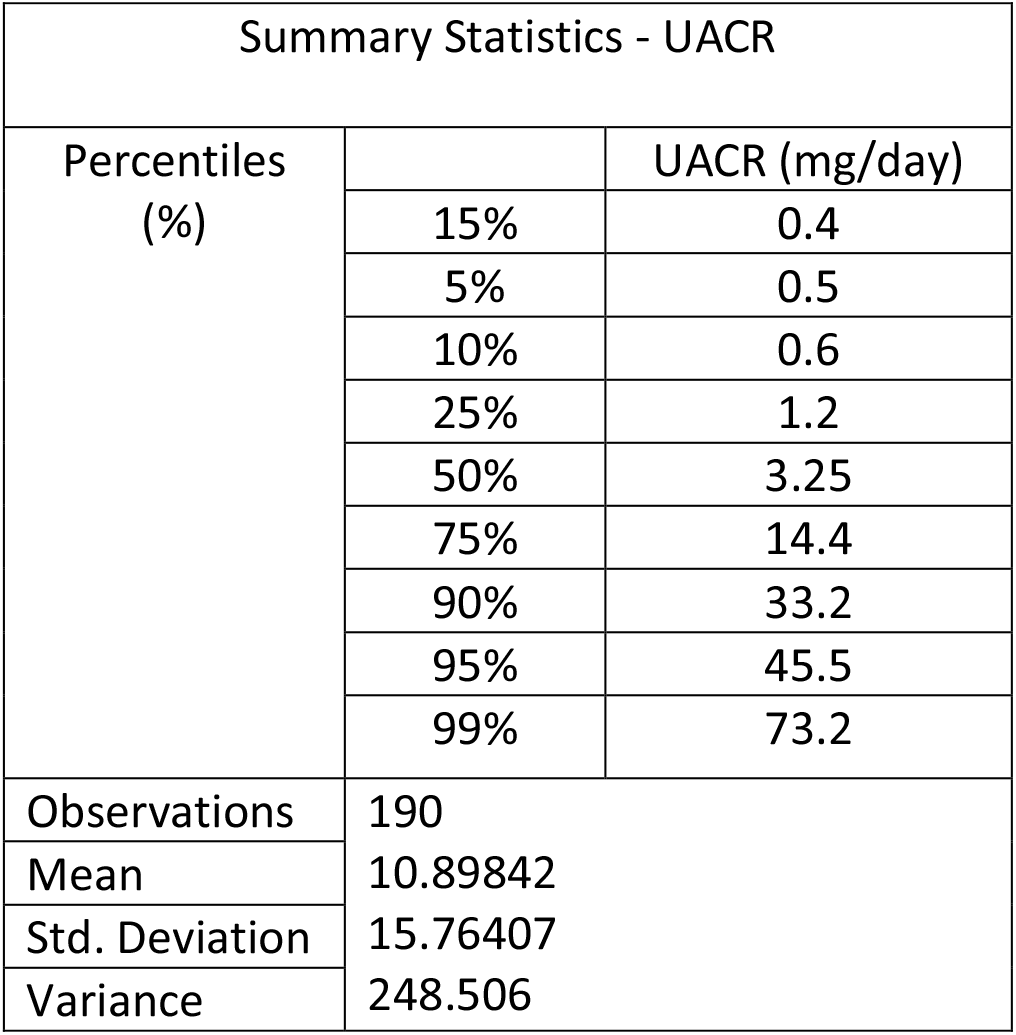
Summary statistics - UACR.

**Figure 3.**
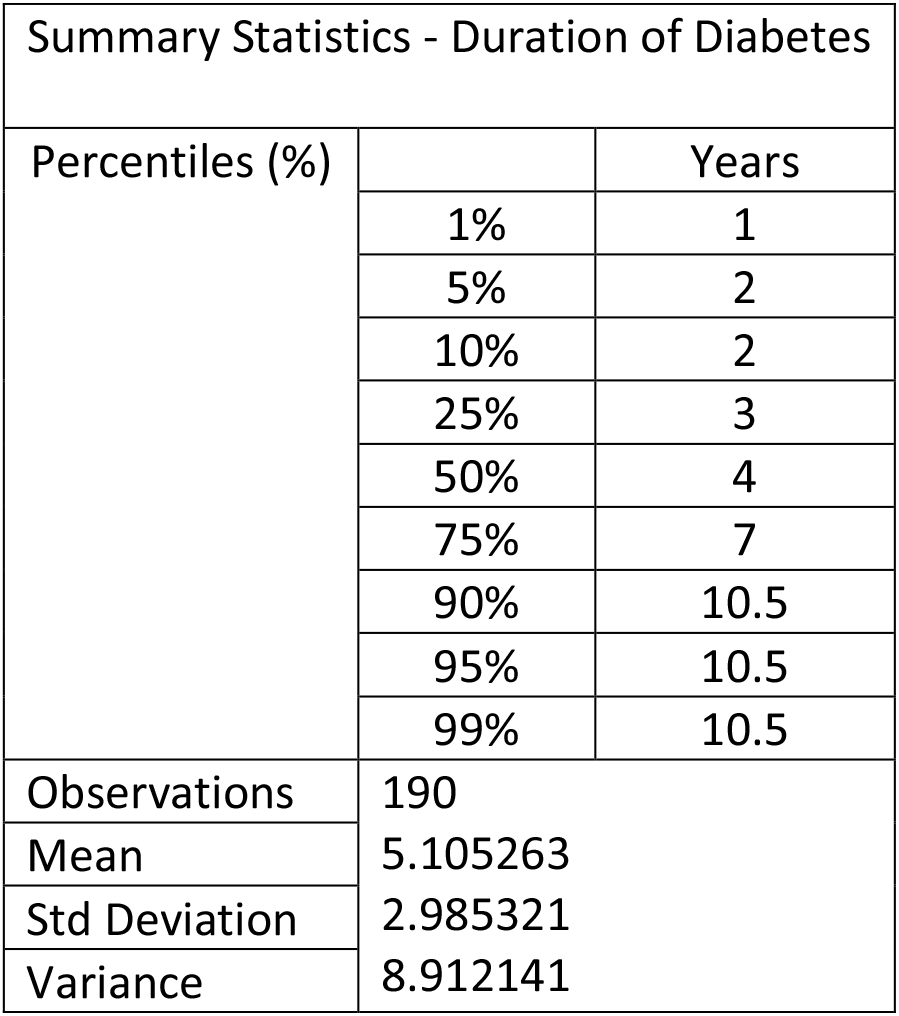
Summary statistics – Duration of Diabetes.

## Discussion

The mean HbA_1_C observed in this study was 8.6% with a standard deviation (SD) of 2.6 and a relatively smaller variance of 6.64 compared to UACR with a mean of 10.9 mg/day, SD of *15*.*76* and variance of *248*.*51*. 90% of the UACR values were below microalbuminuria threshold (< 33.2mg/day) whilst 90% of HbA_1_C readings were above optimal level (5.8%). This showed that 90% of the participants had poor glycemic control in the preceding 3-4 months, indicating their predisposition to development of micro- and macro-vascular complications, including microalbuminuria and diabetic nephropathy, if such has been or will be the state of glycemic control for longer than the assessed period. 90% of UACR < 33.2 mg/day indicated that most participants were normoalbuminuric at the time of testing, with a low mean of 10.9mg/day but a high dispersion of individual values from the mean as demonstrated by a significantly large SD (15.76) and variance (248.51). Normoalbuminuric status in patients with diabetes of many years may not necessarily exclude presence or progression of diabetic nephropathy as indicated by recent evidence suggesting possible independent non-albuminuric pathways for development of kidney disease in diabetics^17-19^.

There was a positive linear relationship between HbA_1_C and UACR readings with fewer data points above the microalbuminuria level (>30 mg/day) [graphs 1&2]. The regression model [figure 4] indicated a significant (P=0.02), though weak (R^2^=0.028 <0.70), positive correlation between UACR and HbA_1_C at 95% statistical confidence. Only 2.8% (R^2^=0.028) of the change in UACR was accounted for by HbA_1_C values (95% statistical confidence). This association between UACR and HbA_1_C was marginally significant (p=0.056) after controlling for duration of illness, weight, systolic blood pressure, systolic hypertension, gender and age in the multivariate regression [figure 5]. Current evidence also supports an association between microalbuminuria and HbA_1_C variability or high persistence^7-8^.

**Graph 1.**
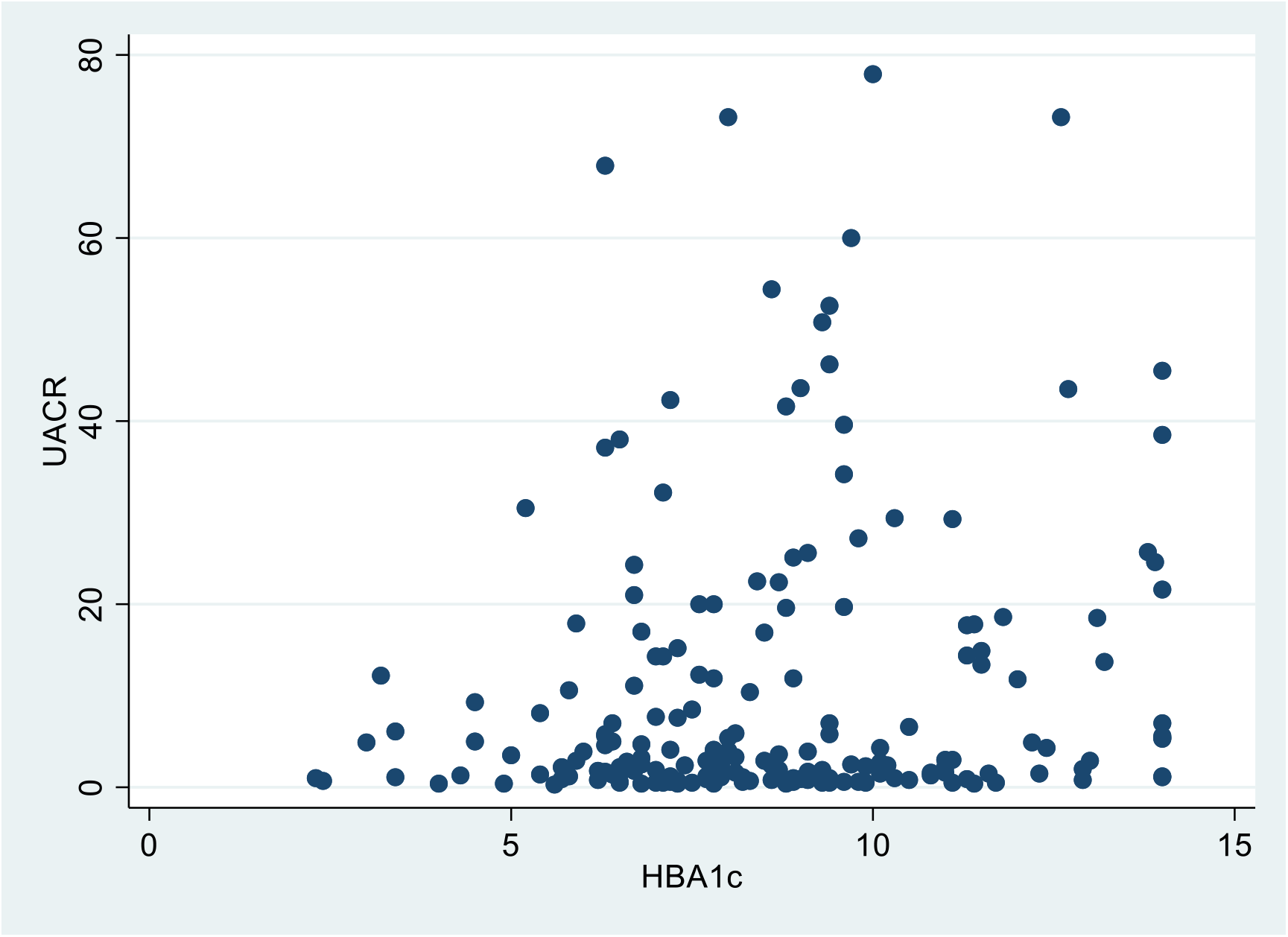
Scatter plot showing relationship between UACR and HbA_1_C.

**Graph 2.**
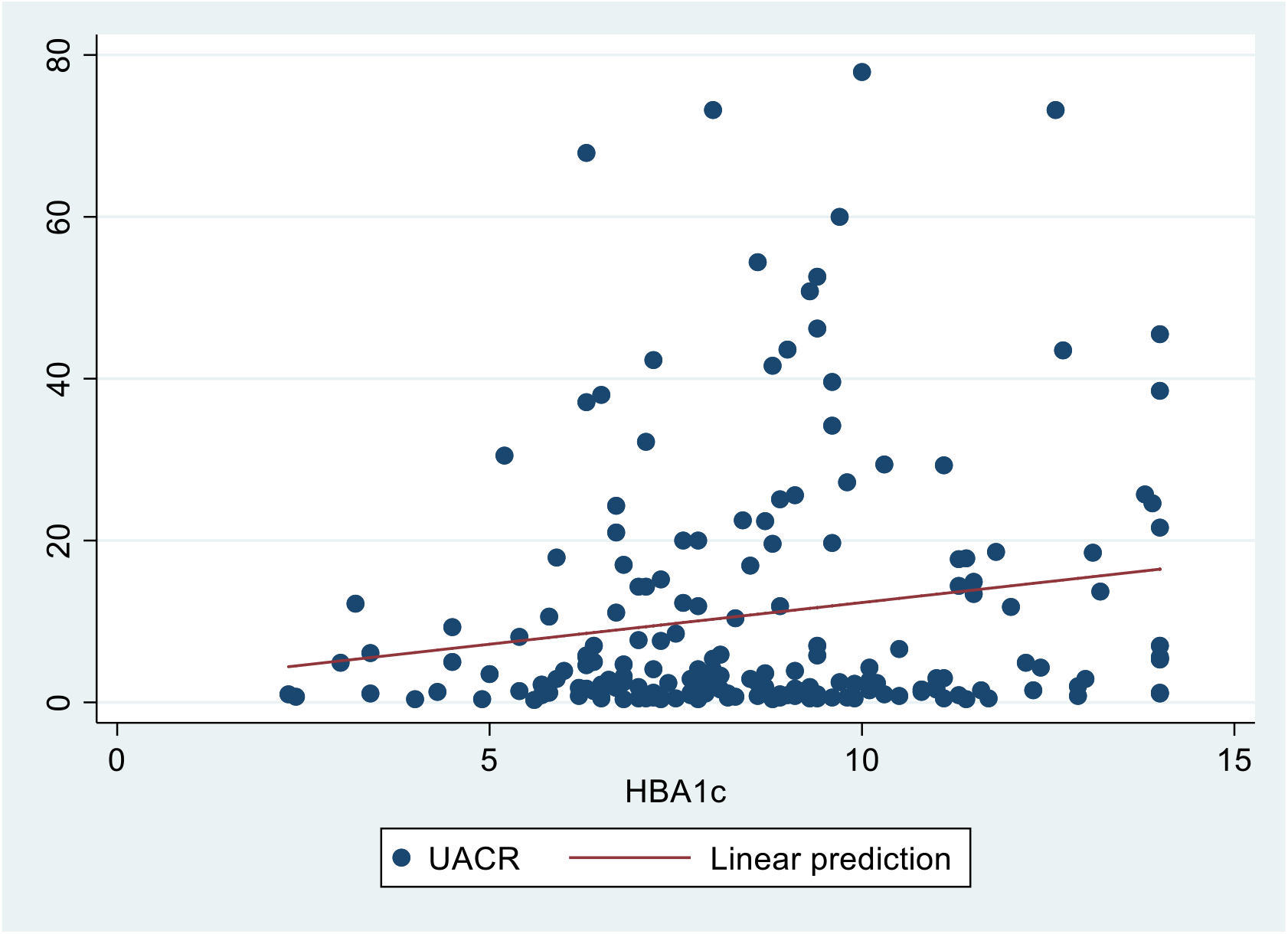
Scatter plot with best-fit line showing predicted linear relationship between UACR and HbA_1_C. • Positive, though weak (R^2^=0.028 <0.70), Correlation between single point UACR and HbA_1_C levels.

**Figure 4.**
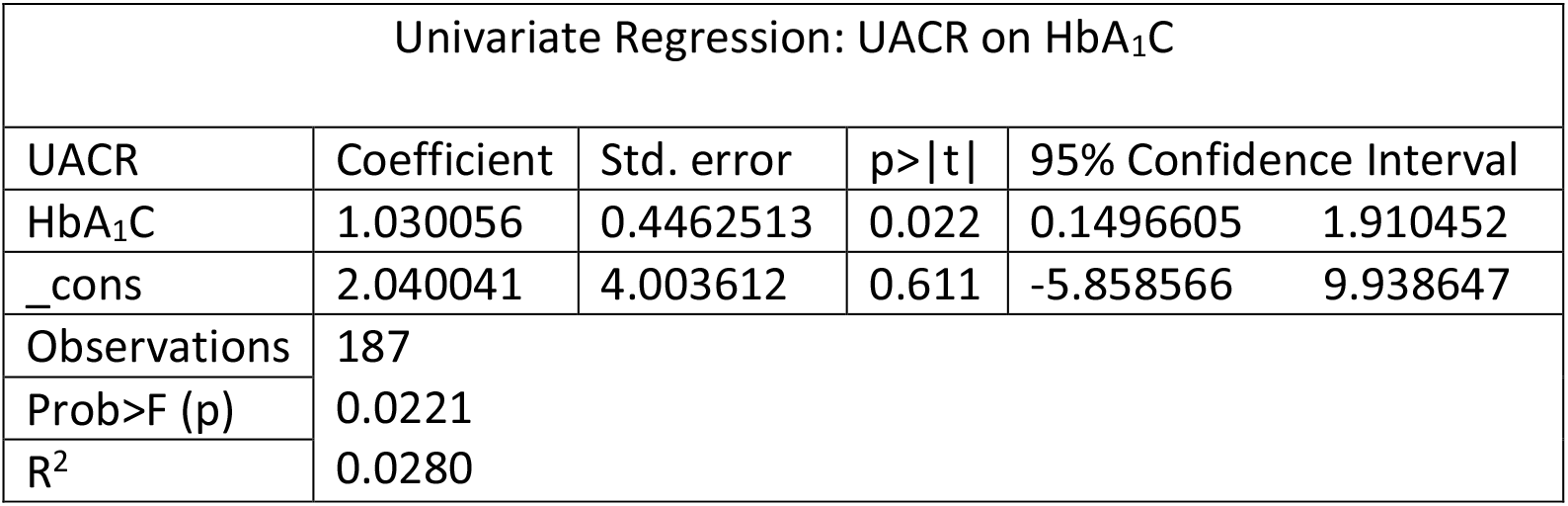
Linear regression output for UACR on HbA_1_C. • Significant (p=0.022), though weak (R^2^=0.028 <0.70), correlation between UACR and HbA_1_C. • Only 2.8% (R^2^=0.0280) of the change in UACR is being accounted for by HbA_1_C with 95% confidence. • A unit increase in HbA_1_C will cause a 1.03 (coef. hba1c=1.030056) increase in UACR [95% statistical confidence].

**Figure 5.**
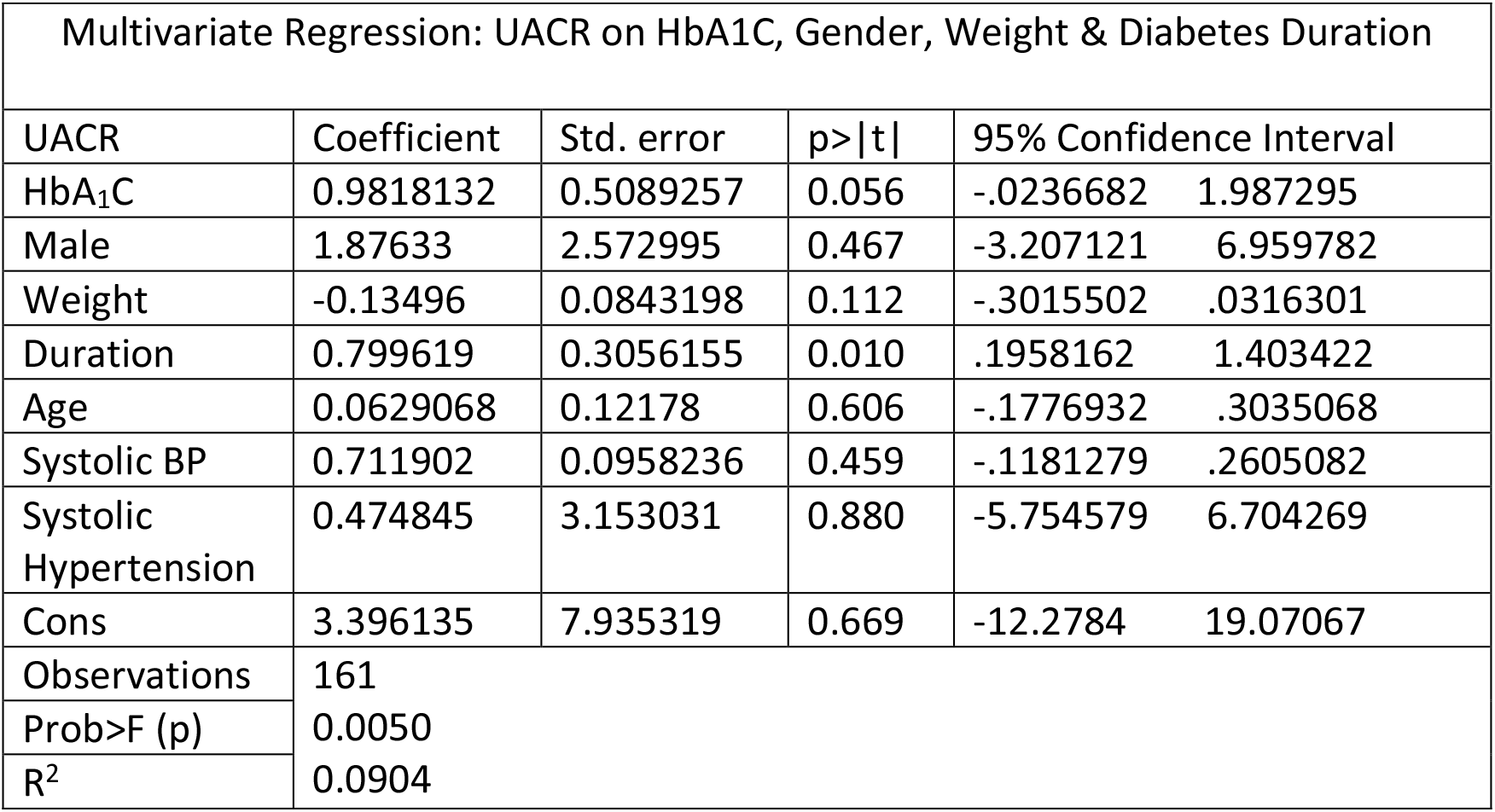
Multivariate Linear Regression: UACR on weight, systolic BP, HbA_1_C, gender, age, and duration of diabetes. • Marginally significant (p=0.056) and weak (R^2^=0.09<0.70) correlation between UACR and HbA_1_C in multivariate regression, controlling for weight, systolic BP, gender, duration of diabetes, age and systolic hypertension. • Significant (p=0.01), though weak (R^2^=0.09<0.70), correlation between duration of diabetes and HbA_1_C in multivariate regression, controlling for potential confounders. • A unit increase in duration of diabetes will cause a 0.8 (coef.duration =0.799619) increase in UACR with 95% statistical confidence. • Overall multivariate model showed significant (p value = 0.0050, f test) but weak association (R^2^=0.0904<0.70): only 9% of the change in UACR is being accounted for by any of the regressed variables individually.

The low prevalence of microalbuminuria amongst study participants, with high HbA_1_C levels, and weakness of association between UACR and HbA_1_C, imply that microalbuminuria may not be singularly influenced by high or variable HbA_1_C levels despite evidence of possible shared or related biochemical mechanisms in their developments^9-14^. Literature also suggest a multifactorial causality in the development of microalbuminuria and diabetic kidney disease, including glomerular hyperfiltration, oxidative stress, hypertension, inflammatory mediators and collagen alteration alongside endothelial dysfunction^12, 20-23^. A High HbA_1_C level, initial or progressive, may therefore not be very useful as an independent predictor of microalbuminuria and diabetic nephropathy in adult diabetics with at least 1-year disease duration. However, in resource-constrained settings without the capacity for microalbuminuria testing, high or variable HbA1c levels can still be used as a proxy to suspecting presence or progression of microalbuminuria, given the linear association and supporting evidence, to prompt appropriate and timely interventions to treat or prevent further development of diabetic nephropathy, as well as related micro- and macro-vascular complications^15^. More studies with larger sample sizes are necessary to further ascertain the strength of the identified association between UACR and high HbA1C levels.

The mean duration of diabetes was 5.11 years with SD=2.99 and 50% of participants at less than 4 years disease duration. This may explain the observed low prevalence of microalbuminuria amongst participants studied as half of them were recently diagnosed. However, related studies have shown presence of microalbuminuria in diabetics in as early as 1-3 years with prevalence increasing with increasing duration^24,25^. Time of diagnosis in our Melanesian subjects may not necessarily reflect duration of illness, as delayed presentation and/or diagnosis are common in clinical practices in the developing world, given logistic and resource limitations coupled with unfavorable patient behavior that often limit early diagnosis^26^. The disease may have progressed undetected to variable stages with subclinical or overt complications at the time of a definitive diagnosis. Therefore, a high index of suspicion for microalbuminuria and diabetic nephropathy, as well as related vascular complications, is necessary even at the time of diagnosis or in recently diagnosed diabetics. A follow-up study involving a cohort of newly diagnosed diabetics is necessary to assess time-to-event outcomes for microalbuminuria in our Melanesian population and compare to studies from other populations.

A significant (P=0.002, CI 0.319 – 1.372), though weak (R^2^=0.0507), correlation between duration of diabetes and UACR (95% statistical confidence) was observed in the linear regression model [figure 6], which remained significant (p=0.01) after controlling for confounders in the multivariate regression [figure 5]. A unit increase in duration of diabetes would entail a 0.85 (coef=0.8459958) increase in UACR. This outcome agrees with existing evidence in support of a strong association between diabetic complications and duration of illness^27-30^, and further highlights the need for heightened clinical suspicion with timely action for diagnosis, treatment and prevention of diabetic complications at earlier stages to reduce morbidity and death.

**Figure 6.**
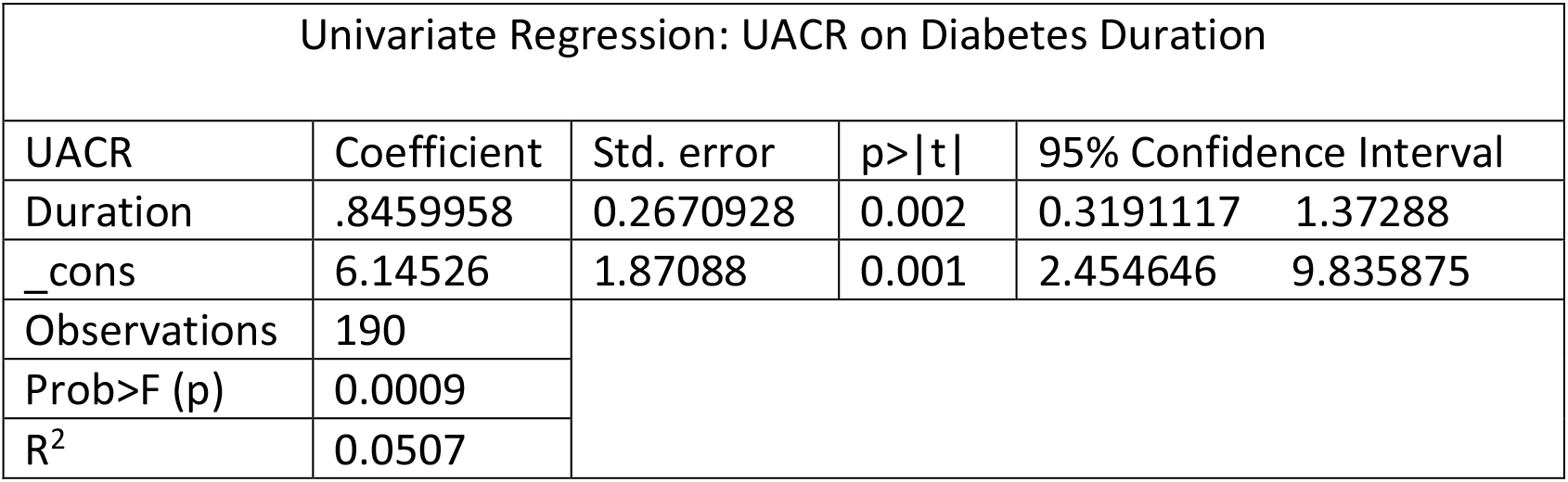
Linear regression UACR on Duration of Diabetes. • Significant (P=0.002, CI 0.319 – 1.372), though weak (R^2^=0.0507 <0.70), correlation between duration of diabetes and UACR with 95% statistical confidence. • A unit increase in diabetes duration will cause a 0.85 (coef.duration =0.8459958) increase in UACR with 95% statistical confidence.

There was no significant association (p=0.880) between single-point systolic hypertension and UACR levels on the multivariate regression, which highlights the fact that single blood pressure readings do not reliably reflect long-term blood pressure status, and therefore may not necessarily represent the effect of long-term blood pressure variability on microalbuminuria development or independent renovascular disease, as suggested by literature on the association of hypertension and microalbuminuria or kidney disease^31-33^.

## Conclusion

A significant (p=0.022), though weak, positive correlation was observed between UACR and HbA1C levels, which may limit the value of HbA_1_C as a reliable, independent predictor for the presence or progression of microalbuminuria and diabetic kidney disease. However, in settings without microalbuminuria testing, high or variable HbA1c levels can be used as a proxy to heighten clinical suspicion for microalbuminuria in order to prompt appropriate and timely interventions to prevent onset or progression of diabetic nephropathy, and related micro-and macro-vascular diabetic complications.

A significant (p=0.002) positive correlation between duration of diabetes and microalbuminuria observed emphasizes the need for heightened suspicion for presence or progression of diabetic nephropathy in patients with diabetes of many (≥5) years. A High prevalence of undiagnosed diabetes with subclinical onset and progression of vascular complications amongst local Melanesian populations further highlights the need for anticipation of microalbuminuria and diabetic nephropathy even at the time of diagnosis.

## Abbreviations

HbA_1_C or A_1_C: Hemoglobin A_1_C
UACR: Urine Albumin-Creatinine Ratio
PNG: Papua New Guinea
R^2^: egression Coefficient

## Ethics

Ethical consideration and approval was granted by the Ethics Review Committee of the Angau Memorial Hospital.

## Acknowledgement

The authors would like to acknowledge the contribution of Professor Isi. H Kevau (FRACP, MMed, Cardiologist-Port Moresby General Hospital), Dr. David Linge (PhD, MMed-Specialist Physician, Port Moresby General Hospital) and Dr. Cathy Timothy (MMed, MBBS-Specialist Physician, Angau Memorial Hospital) without whose invaluable guidance and help this study would not have been possible.

## Reference

1. Persson, F., & Rossing, P. (2018). Diagnosis of diabetic kidney disease: state of the art and future perspective. Kidney international supplements, 8(1), 2–7.

2. Valdez-González, L. A., Méndez-Padrón, A., Gómez-Díaz, R. A., Valladares-Salgado, A., Sánchez-Becerra, M. C., Mondragón-González, R., … Wacher, N. H. (2018). Agreement between the ‘point of care’tests for microalbuminuria and HbA1c performed in mexican family medicine units and the results of standard laboratory tests. Scandinavian journal of clinical and laboratory investigation, 78(1-2), 87–93.

3. Chen, H., Zheng, Z., Huang, Y., Guo, K., Lu, J., Zhang, L., … Jia, W. (2012). A microalbuminuria threshold to predict the risk for the development of diabetic retinopathy in type 2 diabetes mellitus patients. PloS one, 7(5), e36718.

4. Bello, A. K., Levin, A., Tonelli, M., Okpechi, I. G., Feehally, J., Harris, D., … Qarni, B. (2017). Assessment of global kidney health care status. Jama, 317(18), 1864–1881.

5. Burtis, C. A., Ashwood, E. R., & Bruns, D. E. (2012). Tietz textbook of clinical chemistry and molecular diagnostics-e-book. Elsevier Health Sciences.

6. World Health Organization. (2011). Use of glycated haemoglobin (HbA1c) in diagnosis of diabetes mellitus: abbreviated report of a WHO consultation (No. WHO/NMH/CHP/CPM/11.1). Geneva: World Health Organization.

7. Hsu, C. C., Chang, H. Y., Huang, M. C., Hwang, S. J., Yang, Y. C., Lee, Y. S., … Tai, T. Y. (2012). HbA 1c variability is associated with microalbuminuria development in type 2 diabetes: a 7-year prospective cohort study. Diabetologia, 55(12), 3163–3172.

8. Dorajoo, S. R., Ng, J. S. L., Goh, J. H. F., Lim, S. C., Yap, C. W., Chan, A., & Lee, J. Y. C. (2017). HbA1c variability in type 2 diabetes is associated with the occurrence of new-onset albuminuria within three years. Diabetes research and clinical practice, 128, 32–39.

9. Deckert, T., Feldt-Rasmussen, B., Borch-Johnsen, K., Jensen, T., & Kofoed-Enevoldsen, A. (1989). Albuminuria reflects widespread vascular damage. Diabetologia, 32(4), 219–226.

10. Nieuwdorp, M., Mooij, H. L., Kroon, J., Atasever, B., Spaan, J. A., Ince, C., … Kastelein, J. J. (2006). Endothelial glycocalyx damage coincides with microalbuminuria in type 1 diabetes. Diabetes, 55(4), 1127–1132.

11. Parving, H. H. (1996). Microalbuminuria in essential hypertension and diabetes mellitus. Journal of hypertension. Supplement: official journal of the International Society of Hypertension, 14(2), S89–93.

12. Satchell, S. C., & Tooke, J. E. (2008). What is the mechanism of microalbuminuria in diabetes: a role for the glomerular endothelium? Diabetologia, 51(5), 714.

13. Okado, A., Kawasaki, Y., Hasuike, Y., Takahashi, M., Teshima, T., Fujii, J., & Taniguchi, N. (1996). Induction of apoptotic cell death by methylglyoxal and 3-deoxyglucosone in macrophage-derived cell lines. Biochemical and biophysical research communications, 225(1), 219–224.

14. Stehouwer, C. D., Lambert, J., Donker, A. J. M., & van Hinsbergh, V. W. (1997). Endothelial dysfunction and pathogenesis of diabetic angiopathy. Cardiovascular research, 34(1), 55–68.

15. Cheng, D., Fei, Y., Liu, Y., Li, J., Xue, Q., Wang, X., & Wang, N. (2014). HbA1C variability and the risk of renal status progression in diabetes mellitus: a meta-analysis. PLoS One, 9(12), e115509.

16. Oellgaard, J., Gæde, P., Rossing, P., Persson, F., Parving, H. H., & Pedersen, O. (2017). Intensified multifactorial intervention in type 2 diabetics with microalbuminuria leads to long-term renal benefits. Kidney international, 91(4), 982–988.

17. MacIsaac, R. J., Tsalamandris, C., Panagiotopoulos, S., Smith, T. J., McNeil, K. J., & Jerums, G. (2004). Nonalbuminuric renal insufficiency in type 2 diabetes. Diabetes care, 27(1), 195–200.

18. Tsalamandris, C., Allen, T. J., Gilbert, R. E., Sinha, A., Panagiotopoulos, S., Cooper, M. E., & Jerums, G. (1994). Progressive decline in renal function in diabetic patients with and without albuminuria. Diabetes, 43(5), 649–655.

19. Caramori, M. L., Fioretto, P., & Mauer, M. (2003). Low glomerular filtration rate in normoalbuminuric type 1 diabetic patients: an indicator of more advanced glomerular lesions. Diabetes, 52(4), 1036–1040.

20. Campos-Pastor, M. M., Escobar-Jimenez, F., Mezquita, P., Herrera-Pombo, J. L., Hawkins-Carranza, F., Luna, J. D., … Rigopoulos, M. (2000). Factors associated with microalbuminuria in type 1 diabetes mellitus: a crosssectional study. Diabetes research and clinical practice, 48(1), 43–49.

21. Chen, S., Hong, S. W., Iglesias-dela Cruz, M. C., Isono, M., Casaretto, A., & Ziyadeh, F. N. (2001). The key role of the transforming growth factor-β system in the pathogenesis of diabetic nephropathy. Renal failure, 23(3-4), 471–481.

22. Wolf, G., Chen, S., & Ziyadeh, F. N. (2005). From the periphery of the glomerular capillary wall toward the center of disease: podocyte injury comes of age in diabetic nephropathy. Diabetes, 54(6), 1626–1634.

23. Jensen, J. S., Feldt-Rasmussen, B., Strandgaard, S., Schroll, M., & Borch-Johnsen, K. (2000). Arterial hypertension, microalbuminuria, and risk of ischemic heart disease. Hypertension, 35(4), 898–903.

24. Warram, J. H., Gearin, G., Laffel, L., & Krolewski, A. S. (1996). Effect of duration of type I diabetes on the prevalence of stages of diabetic nephropathy defined by urinary albumin/creatinine ratio. Journal of the American Society of Nephrology, 7(6), 930–937.

25. Mogensen, C. E., & Poulsen, P. L. (1994). Epidemiology of microalbuminuria in diabetes and in the background population. Current opinion in nephrology and hypertension, 3(3), 248–256.

26. Venkataraman, K., Kannan, A. T., & Mohan, V. (2009). Challenges in diabetes management with reference to India. International journal of diabetes in developing countries, 29(3), 103.

27. Ahmed, U. (2004). Prevalence of chronic complications and associated factors in type 2 diabetes. J Pak Med Assoc, 54, 54–59.

28. Orchard, T. J., Dorman, J. S., Maser, R. E., Becker, D. J., Drash, A. L., Ellis, D., … Kuller, L. H. (1990). Prevalence of complications in IDDM by sex and duration: Pittsburgh Epidemiology of Diabetes Complications Study II. Diabetes, 39(9), 1116–1124.

29. Molitch, M. E., DeFronzo, R. A., Franz, M. J., & Keane, W. F. (2004). Nephropathy in diabetes. Diabetes care, 27, S79.

30. Molitch, M. E., DeFronzo, R. A., Franz, M. J., & Keane, W. F. (2004). Nephropathy in diabetes. Diabetes care, 27, S79.

31. Cirillo, M., Stellato, D., Laurenzi, M., Panarelli, W., Zanchetti, A., & De Santo, N. G. (2000). Pulse pressure and isolated systolic hypertension: association with microalbuminuria. Kidney international, 58(3), 1211–1218.

32. Høegholm, A., Bang, L. E., Kristensen, K. S., Nielsen, J. W., & Holm, J. (1994). Microalbuminuria in 411 untreated individuals with established hypertension, white coat hypertension, and normotension. Hypertension, 24(1), 101–105.

33. Parving, H. H., Mogensen, C. E., & Evrin, P. E. (1974). Increased urinary albumin-excretion rate in benign essential hypertension. The Lancet, 303(7868), 1190–1192.

